# Reporting of paediatric exercise-induced respiratory symptoms by physicians and parents

**DOI:** 10.1101/2024.04.30.24306617

**Authors:** Eva SL Pedersen, Sarah Glick, Carmen CM de Jong, Cristina Ardura-Garcia, Anja Jochmann, Carmen Casaulta, Katharina Hartog, Diana Marangu-Boore, Dominik Mueller-Suter, Nicolas Regamey, Florian Singer, Alexander Moeller, Claudia E Kuehni

**Affiliations:** Institute of Social and Preventive Medicine, University of Bern, Switzerland; Department of Clinical Research, University of Bern, Switzerland; Paediatric Respiratory Medicine, Children’s University Hospital of Bern, University of Bern, Switzerland; Department of Paediatrics, University Children’s Hospital, University of Basel, Switzerland; Division of Paediatric Pulmonology, Children’s Hospital Chur, Chur, Switzerland; Division of Paediatric Pulmonology, Children’s Hospital St. Gallen, Switzerland; Department of Paediatrics and Child Health, University of Nairobi, Nairobi, Kenya; Department of Paediatrics, Kantonsspital Aarau, Switzerland; Division of Paediatric Pulmonology, Children’s Hospital of Central Switzerland, Lucerne, Switzerland; Department of Paediatric Pulmonology, University Children’s Hospital Zurich, University of Zurich, Switzerland; Division of Paediatric Pulmonology and Allergology, Department of Paediatrics and Adolescent Medicine, Medical University of Graz, Graz, Austria

**Keywords:** exercise-induced, asthma, dysfunctional breathing, respiratory symptoms, reporting, agreement

## Abstract

**Aims of the study:** Routinely collected health data are increasingly used for research, however important history items may be incomplete in medical records. We assessed clinical documentation of exercise-induced respiratory symptoms (EIS) by treating physicians and compared with parent-reported EIS for the same children.

**Methods:** We analysed data from the Swiss Paediatric Airway Cohort (SPAC), a multicentre observational study of children treated in Swiss outpatient pulmonology clinics. We included children 6 to 17 years of age who were referred to a paediatric pulmonologist for evaluation of EIS. Features of EIS recorded by physicians were extracted from outpatient clinical letters transmitted to the referring physician, while parent-reported EIS data were collected from a standardized questionnaire completed at SPAC enrolment. We calculated agreement between physician-documented and parent-reported EIS characteristics using Cohen’s and Fleiss’s kappa.

**Results:** Of 1669 children participating in SPAC (2017-2019), 193 (12%) met the inclusion criteria, of whom 48% were girls. Physicians provided detailed information on EIS in 186 (96%) outpatient clinical letters. Documented characteristics included: type of physical activity triggering EIS (69%), localisation of EIS in chest or throat (48%), respiratory phase of EIS (45%), and timing of EIS during or after exercise (37%). Previous bronchodilator use (94%) and its effect on EIS (88%) were consistently documented by physicians. The clinical letters of children diagnosed with dysfunctional breathing more often contained detailed EIS characteristics than for children diagnosed with asthma. The agreement between physician-documented and parent-reported EIS was moderate for use of bronchodilators (k=0.53) and poor to fair for all other features (k=0.01-0.36).

**Conclusion:** This study highlights that outpatient clinical letters may lack some details on EIS characteristics, information which parents could provide. A standardized and detailed method for documenting paediatric respiratory symptoms in the coordinated data infrastructure may enhance future analyses of routinely collected health data.

## Introduction

Exercise-induced respiratory symptoms (EIS) are common in childhood (1). Symptom characteristics vary depending on the underlying respiratory aetiology (2-4), which may include asthma, dysfunctional breathing (e.g. inducible laryngeal obstruction, breathing pattern disorders), or insufficient fitness (5-9). The EIS domains – such as timing of onset, severity, perceived localisation, relation to exercise intensity and duration, and response to treatment are important features in the clinical history. Exercise-induced wheeze, cough, and chest tightness are common in asthma (10, 11), while stridor and dyspnoea are more often noted for extrathoracic dysfunctional breathing (9, 11). Thoracic dysfunctional breathing is associated with exertional dyspnoea, sighing, and dizziness, and may be accompanied by hyperventilation (12). Poor differentiation between isolated or co-existing dysfunctional breathing and other entities may lead to diagnosis misclassification, overtreatment (e.g., inhaled corticosteroids) and excess health care costs (13, 14). Thus, detailed history-taking is essential for evaluating the differential diagnosis in children referred with EIS.

With the advent of electronic health record databases and the creation of clinical data repositories, also known as coordinated data infrastructures, secondary use of routinely collected health data to promote public health research is evolving (15). The sharing of depersonalized health data will be increasingly important for enhancing the understanding, management, and prevention of disease (16). The Swiss Personalized Health Network (SPHN) aims to improve the utilization of data obtained during routine healthcare encounters by facilitating accessibility for research (17). Swiss paediatricians agreed to uniformly capture a minimal set of variables in medical records (18), but recording of symptoms was not standardized yet.

To our knowledge, the comprehensiveness of EIS documentation in outpatient clinical letters has not been previously studied. This information is important for understanding the usability of EIS-related data for healthcare research through, for example, text search or similar methods. We analyzed the characteristics of EIS documentation in outpatient clinical letters and assessed the agreement with parental information from a standardized questionnaire.

## Materials and methods

### Study design

We used data from the Swiss Paediatric Airway Cohort (SPAC), a national multicentre longitudinal study of children evaluated in outpatient paediatric pulmonology clinics throughout Switzerland (clinicaltrials.gov: nr. NCT03505216) (19-22). The SPAC study aims to enrol all children referred to the pulmonology clinics for respiratory problems such as wheeze, cough, dyspnoea, and exercise-related symptoms. The SPAC study is observational and embedded in routine medical care, and all diagnostic tests are performed per clinical indication. Recruitment started in July 2017 and is ongoing.

In conjunction with SPAC study enrolment, parents complete a baseline questionnaire, which collects data related to respiratory symptoms, medication utilization, environmental exposures, and health behaviours at the time of referral. We also collect data from medical records, including reason for referral, diagnostic investigations, final diagnosis, and prescribed treatment. All data are entered into a Research Electronic Data Capture (REDCap) database (23). Written informed consent is obtained at enrolment from parents and directly from participants older than 13 years. The SPAC study is approved by the Bern Cantonal Ethics Committee (Kantonale Ethikkomission Bern 2016-02176).

### Inclusion criteria

We included children aged 6-17 years who enrolled in the SPAC study between July 1, 2017 and December 1, 2019, and who had been primarily referred to the paediatric pulmonology clinic for EIS. We considered EIS the main reason for referral when the referral letter or the first outpatient clinical letter listed EIS as the sole or main reason for specialist consultation.

### Physician-documented and parent-reported EIS data

We obtained physician-documented EIS data from the outpatient clinical letter that was transmitted to the referring physician. Physicians’ notes were individually reviewed by a SPAC investigator (EP), and relevant data was manually extracted from the text. Extracted data included: type of symptom experienced by the child (e.g., wheeze, cough, dyspnoea, tingling feeling in fingertips or lips); perceived symptom localisation (chest or throat); respiratory phase in which symptoms occurred or were felt maximally (inspiration or expiration); triggers of EIS (specific physical activities); timing of onset of EIS (during or after exercise); and use and effect of bronchodilator treatment on EIS (**Table 1**). Parent-reported EIS data was extracted from the baseline questionnaire (**Table 1**).

**Table 1.** Information extracted from outpatient clinical letter (physician) and standardized questionnaire (parent) to measure agreement in reporting EIS characteristics.

| Variable | Physician documented EIS, extracted from outpatient clinic letter | Question in questionnaire |
| --- | --- | --- |
| **Any EIS** | Does the child have any exercise-induced symptoms (yes, no, not mentioned) | Does your child sometimes experience breathing problems when exercising? (yes/no) |
| **Wheeze** | Expiratory wheeze | Which breathing problems does your child experience when exercising? Wheezing or whistling breathing sounds (yes/no) |
| **Cough** | Cough | Which breathing problems does your child experience when exercising? Cough (yes/no) |
| **Dyspnoea** | Dyspnoea, shortness of breathing, difficulty breathing | Which breathing problems does your child experience when exercising? Dyspnoea or tightness (yes/no) |
| **Tingling feeling in fingertips or lips** | Tingling feeling in fingertips or lips | Does your child sometimes experience a sensation as if ants were creeping around the fingertips or lips while exercising? (yes/no) |
| **Other** |  | Which breathing problems does your child experience when exercising? Other problems (describe) |
| **Types of EIS triggers** | Which of the following activities trigger exercise-induced symptoms (running, bicycle riding, intensive sport games, swimming, cold weather sports, no exercise triggers specified in letter) | In which of the following situation does the breathing problems occur? Running short, middle, longer distances; biking; sport games; swimming |
| **Localisation of dyspnoea** | Where are the exercise-induced symptoms mainly felt (chest, throat, chest and throat, not specified in letter) | If dyspnoea or tightness, where is the sensation felt the strongest? (Chest, throat, everywhere (chest and throat)) |
| **When are EIS worst** | In which respiratory phase are the exercise-induced symptoms worst (inspiration, expiration, inspiration and expiration, not specified in letter) | When are the breathing problems worst? (during inspiration, during expiration, equally during inspiration and expiration) |
| **EIS start** | When do the exercise-induced symptoms begin (during exercise, after exercise, not specified) | When do the breathing problems begin? – When you child for example runs a longer distance, then the breathing problems normally begin: (choose one) straight away after the first steps, a few minutes after beginning the exercise, after ending the exercise |
| **Inhalation for EIS** | Has the child used any inhaled medication before exercise (yes, no, not mentioned) | Has your child inhaled an asthma-spray or asthma-powder for breathing problems occurring when exercising? (yes/no) |
| **Effect of inhalation** | How well does the medication help (symptoms disappear, symptoms are reduced, no difference, not mentioned) | If yes: How well does the medication help? (choose one): the breathing problems disappear, the breathing problems are reduced, you almost feel no difference |

EIS = exercise-induced respiratory symptom

### Medical diagnosis associated with EIS

The final diagnosis assigned by the paediatric pulmonologist was extracted from the outpatient clinical letter. We grouped diagnoses into five major categories consistent with published literature: 1) asthma, 2) extrathoracic dysfunctional breathing (functional: inducible laryngeal obstruction; structural: e.g. laryngomalacia, tracheomalacia), 3) thoracic dysfunctional breathing (functional: EIS with hyperventilation, breathing pattern disorders), 4) asthma plus dysfunctional breathing (for patients with coexisting diagnoses), and 5) other diagnoses (e.g. insufficient fitness level, chronic cough, or rare pulmonary causes) (6, 8).

### Statistical analysis

We compared proportions of physician-documented and parent-reported EIS using numbers and percentages. We calculated agreement for specific characteristics among children where the characteristic was mentioned in both the outpatient clinical letter and the parental questionnaire – e.g. for onset of symptoms, we calculated agreement for whether symptoms started during or after exercise if the clinical letter included information on the timing of onset. Children where this characteristic was not mentioned in the clinical letter were excluded from this analysis. We calculated kappa statistics using Cohen’s kappa for dichotomous outcomes and Fleiss’ kappa for categorical variables with more than two categories. The kappa was interpreted using Landis and Koch’s criteria: 0.0-0.2 slight agreement, 0.21-0.40 fair agreement, 0.41-0.60 moderate agreement, 0.61-0.80 substantial agreement, and 0.81 – 1.0 almost perfect to perfect agreement (24). We also assessed whether agreement of EIS depended on who filled in the baseline questionnaire (mother, father, child-assisted) using kappa statistics. We used STATA version 14 for statistical analyses.

## Results

Of the 1669 children who participated in the SPAC study within the eligibility period, 193 (12%) met the inclusion criteria (**Table 2**). The mean age was 12 years and 92 (48%) were girls. Most study participants were seen in Luzern (33%) followed by Zurich (25%), Basel (17%), Aarau (12%), Bern (11%), and St. Gallen (2%). The final diagnosis from the paediatric pulmonologist was asthma in 106 patients (55%), extrathoracic dysfunctional breathing in 33 (overall 17%; functional in 30 (16%) and structural in 3 (1%)), thoracic dysfunctional breathing in 21 (11%), asthma plus dysfunctional breathing in 21 (11%), and other diagnoses in 12 (6%).

**Table 2.**
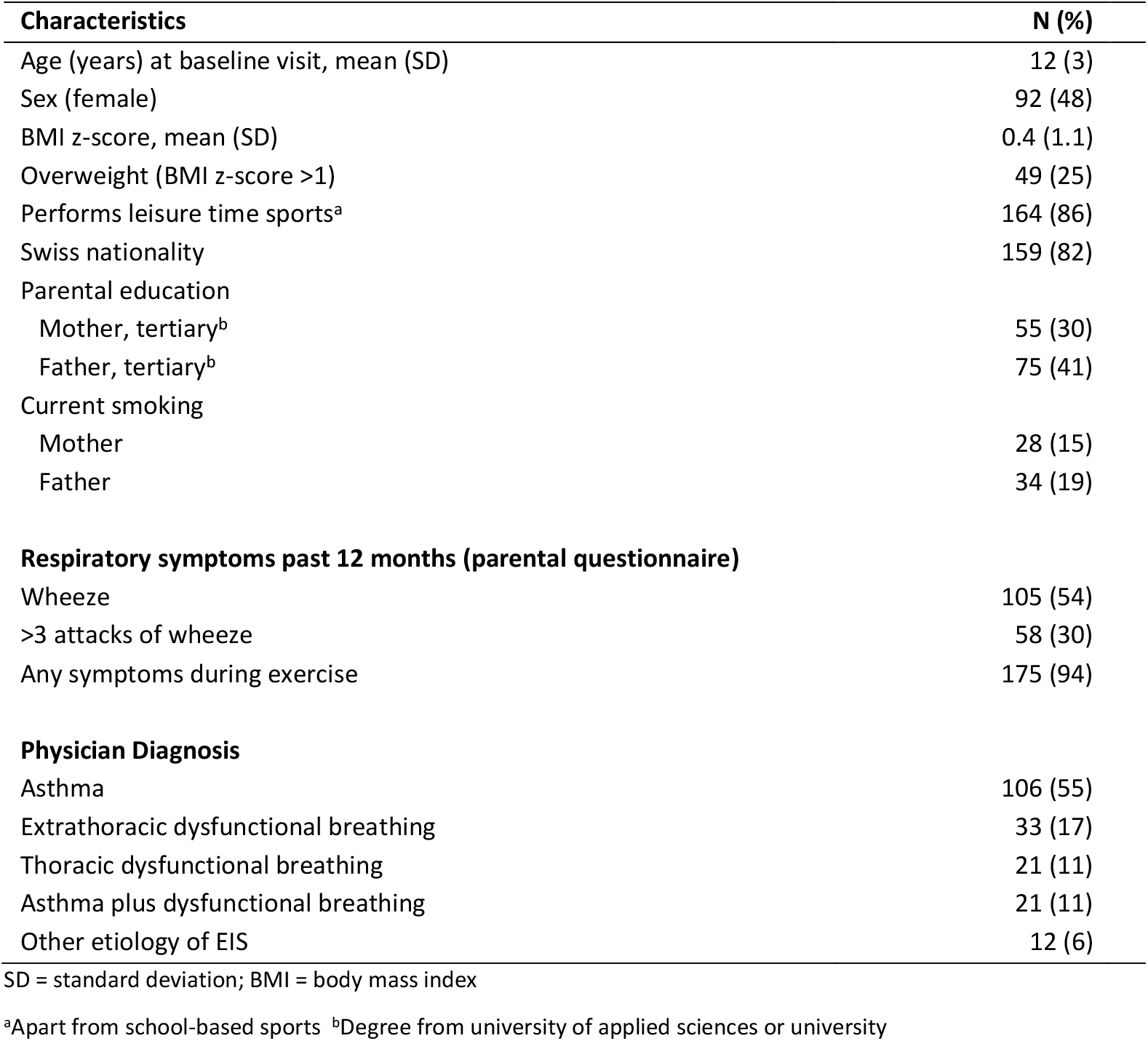
Characteristics of children referred to Swiss pulmonology outpatient clinics for exercise-induced respiratory symptoms (N=193)

The outpatient clinical letter contained characteristics of EIS in 186 (96%) of the 193 children. The type of symptom experienced with exercise was almost always documented by physicians (96%), followed by triggering physical activity (69%), perceived symptom localisation (48%), respiratory phase (45%), and onset of EIS relative to exercise (37%). Bronchodilator use prior to paediatric pulmonology consultation was very frequently documented in the outpatient clinical letter (94%), and the medication effect was noted in 88% of the letters. Overall, characteristics of EIS were more often documented for children diagnosed with dysfunctional breathing than for children diagnosed with asthma or other diagnoses (**Figure 1**).

**Figure 1.**
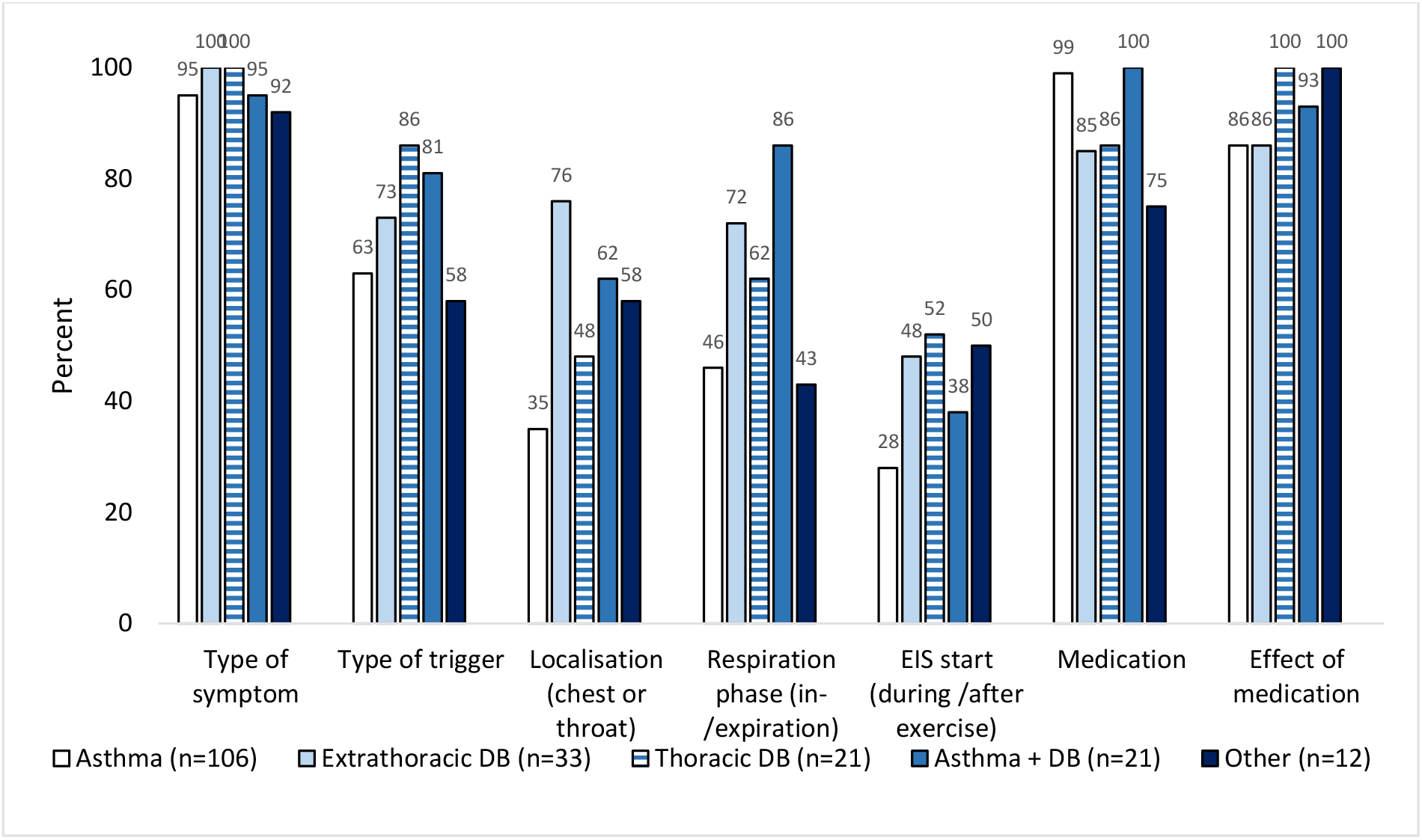
Exercise-induced symptoms documented by the physician in the outpatient clinical letter, by final diagnosis (N=193) EIS = exercise-induced symptoms, DB = dysfunctional breathing

Parents reported the type of respiratory symptom and triggering exercise more often than physicians (**Table 3**). The most frequent symptom, exercise-induced cough, was indicated in 57% of parent-completed questionnaires versus 35% of clinical letters. Similarly, tingling in fingers or lips – the rarest symptom to be noted – was reported more often by parents (18%) than physicians (1%).

**Table 3.** Agreement between physician-documented and parent-reported EIS (N=175)

|  | Physician<br>document<br>ed | Parent<br>reported | Agreement between physician-documented and parent-reported EIS |  |  |  | Kappa |
| --- | --- | --- | --- | --- | --- | --- | --- |
|  |  |  | physician+<br>parent + | physician +<br>parent - | physician -<br>parent + | physician -<br>parent - |  |
| **Type of symptom<sup>a</sup> (n=175)** |  |  |  |  |  |  |  |
| Wheeze or<br>whistling sounds | 56 (32) | 101 (58) | 43 (25) | 13 (7) | 58 (33) | 61 (35) | 0.24 |
| Cough | 61 (35) | 100 (57) | 52 (30) | 9 (5) | 48 (27) | 66 (38) | 0.39 |
| Dyspnoea | 114 (65) | 146 (83) | 101 (58) | 13 (7) | 45 (26) | 16 (9) | 0.17 |
| Tingling in<br>fingers/lips | 2 (1) | 31 (18) | 2 (1) | 0 | 29 (16) | 145 (82) | 0.09 |
| **Type of exercise triggering EIS<sup>b</sup> (n=121)** |  |  |  |  |  |  |  |
| Running | 44 (36) | 75 (62) | 30 (25) | 14 (12) | 45 (37) | 32 (26) | 0.08 |
| Biking | 18 (15) | 79 (65) | 17 (14) | 1 (1) | 62 (51) | 41 (34) | 0.14 |
| Intensive sports | 81 (67) | 94 (78) | 63 (52) | 18 (15) | 31 (26) | 9 (7) | 0.01 |
| Swimming | 8 (7) | 46 (38) | 8 (7) | 0 | 38 (31) | 75 (62) | 0.22 |
| Cold weather sports |  |  |  |  |  |  |  |
| **Symptom localisation (n=73)** |  |  |  |  |  |  | 0.36 |
| Chest | 40 (55) | 41 (56) | * |  |  |  |  |
| Throat | 26 (36) | 12 (16) |  |  |  |  |  |
| Chest and Throat | 7 (10) | 20 (27) |  |  |  |  |  |
| **Respiratory phase<sup>c</sup> (n=94)** |  |  |  |  |  |  | 0.13 |
| Inspiration | 47 (50) | 47 (50) | * |  |  |  |  |
| Expiration | 37 (39) | 5 (5) |  |  |  |  |  |
| Inspiration and<br>Expiration | 10 (11) | 42 (45) |  |  |  |  |  |
| **Timing of EIS onset<sup>e</sup> (n=66)** |  |  |  |  |  |  | 0.19 |
| During exercise | 45 (68) | 55 (83) | 40 (61) | 5 (8) | 15 (23) | 6 (9) |  |
| After exercise | 21 (32) | 11 (17) | 6 (9) | 15 (23) | 5 (8) | 40 (61) |  |
| **Prior use of bronchodilator for EIS<sup>f</sup> (n=159)** |  |  |  |  |  |  | 0.53 |
| Yes | 99 (62) | 111 (70) | 88 (55) | 11 (7) | 23 (14) | 37 (23) |  |
| **Effect of bronchodilator (n=76)** |  |  |  |  |  |  | 0.27 |
| EIS disappears | 23 (30) | 17 (22) | * |  |  |  |  |
| EIS reduced | 29 (38) | 42 (55) |  |  |  |  |  |
| No difference | 24 (32) | 17 (22) |  |  |  |  |  |

We analysed agreement if the characteristic in question was mentioned both in the physician's outpatient clinical letter and the parental questionnaire, thus characteristics that were missing in both documents were excluded from this analysis.
EIS = exercise-induced respiratory symptoms \*Cell percentages cannot meaningfully be displayed because the variable is not dichotomous.

The highest agreement between physicians and parents was found for recording bronchodilator use (k=0.53). The agreement for type of symptom was best for cough (k=0.39) and wheeze (k=0.24) and lowest for tingling in lips or fingers (k=0.09). The agreement for EIS trigger was generally poor across the different categories of exercise (k=0.01-0.14). Perceived localisation of EIS to the chest or throat (k=0.36) had better agreement than respiratory phase of symptoms (k=0.13) and timing of EIS onset (k=0.19). Agreement between physician-documented and parent-reported EIS differed for single items depending on who filled in the questionnaire (mother, father, or child-assisted), but overall agreement was not better for one subgroup.

## Discussion

This study is the first to describe characteristics of EIS documentation by paediatric pulmonologists among children enrolled in a large paediatric respiratory cohort and referred primarily for EIS. Outpatient clinical letters usually included information related to specific symptom(s) and bronchodilator use, but less often provided important additional details such as perceived symptom localisation, respiratory phase, and timing of onset of EIS. These characteristics of EIS have been described as important factors for differentiating dysfunctional breathing from asthma and other aetiologies (3, 11, 25-27), however this information was documented in the outpatient clinical letter in less than half of the children.

There can be several reasons why physicians documented fewer anamnestic details about EIS in the outpatient clinical letter compared to what parents conveyed in the standardized questionnaire. Paediatric pulmonologists may have chosen to document only the most important symptoms in the outpatient clinical report, where succinct summarization is an important consideration for both the writer (treating physician) and reader (referring physician). It’s also possible that physicians verbally inquired about all EIS domains but did not systematically document negative responses in the outpatient clinical report. Another explanation concerns different intrinsic characteristics of the data collection tools. Whereas medical documentation is flexible and unique to each physician, parental questionnaires relied on a standardized checklist of specific symptoms and preformulated response choices.

Interestingly, we observed consistent differences in the extent of EIS documentation by physicians, depending on final diagnosis. When the final diagnosis was extrathoracic or thoracic dysfunctional breathing, all EIS symptom domains (type of symptom, type of trigger, localisation, respiratory phase of symptom, and timing of EIS) were more frequently documented in the outpatient clinical letter versus when the final diagnosis was asthma. We can speculate that since these diagnoses are relatively rarer than asthma, paediatric pulmonologists may have documented more characteristics to communicate differential diagnosis evaluation and clinical decision-making to the referring physician.

Agreement on symptom reporting between physicians and parents varied from poor to moderate. The absence of agreement for specific EIS characteristics was mainly due to parental reporting of symptoms that were not mentioned in the physician letter; only in a few instances did the outpatient clinical letter include symptoms that were not reported in the parental questionnaire. Two studies that investigated agreement between physician documentation and standardized parent reporting of paediatric respiratory symptoms, but which were not specifically focused on EIS, similarly found a poor to fair agreement. A study using population-based data from the Dutch Generation R study found that parents more often reported wheeze than physicians (36% versus 20%, k=0.36), in line with our results (28). Additionally, the WHISTLER Birth Cohort (1007 children, age 5 years) reported a k=0.07 for recent wheeze and a k=0.12 for ever wheezing (29).

The analysis of depersonalized health data from a coordinated data infrastructure will be increasingly important for enhancing the understanding, management, and prevention of disease (16). In Switzerland, several projects within the Swiss Personalized Health Network aim to standardize routine health care data and make it more readily available for research (17, 18). Enhanced symptom documentation could improve the capacity for paediatric respiratory research by bolstering the comprehensiveness of the Swiss data infrastructure. One way to accomplish this would be through inclusion of a standardized checklist of EIS characteristics, to be jointly completed by parents and patients, and stored in the coordinated data infrastructure. Our study suggests that parent questionnaires remain a vital source of information in paediatric research studies and can effectively complement physician documentation, particularly as it pertains to detailed recording of symptom characteristics. This offers a solution that does not oblige paediatric pulmonologists to document EIS with lengthy symptom checklists or preformulated templates, which would lengthen and complicate outpatient clinical letters and not likely enhance physician communication.

This study is strengthened by the real-world, observational design of SPAC, the relatively large size of this cohort, and the inclusion of children from different paediatric respiratory outpatient clinics in Switzerland. Another strength is the inclusion of children with EIS of different aetiologies, thus allowing a comprehensive analysis of EIS documentation which includes both asthma and rarer diagnoses. A limitation of this study is that it only represents children referred to participating paediatric respiratory outpatient clinics in the German language region of Switzerland. We, however, do not have reason to suspect that EIS documentation would differ systematically in the other Swiss regions.

## Conclusion

Characteristics of paediatric EIS are documented differently by physicians and parents, with a generally low level of agreement when comparing the outpatient physician letter with a standardized parent questionnaire. Since comprehensive documentation of paediatric respiratory symptoms is important when analysing data exported from coordinated data infrastructures, a structured way of digitally documenting symptoms could enhance the usability of routine health care data in future paediatric EIS research. Standardized, parent-completed questionnaires should be viewed as an important complement to physician letters when considering the comprehensiveness of the paediatric health data infrastructure.

## Data Availability

All data produced in the present study are available upon reasonable request to the authors

## Acknowledgements

We thank the families who took part in the SPAC study; the research assistants (Natalie Messerli, Gia Thu Ly, Labinata Gjokaj, Meret Ryser, and Malui Frei) for helping with data collection and data entry; PedNet Bern for supporting data collection in Bern; and the members of the SPAC study team. SPAC study team members include D. Mueller-Suter, P. Eng, and B. Kern (Canton Hospital Aarau, Aarau, Switzerland); U. Frey, J. Hammer, A. Jochmann, D. Trachsel, and A. Oettlin (University Children’s Hospital Basel, Basel, Switzerland); P. Latzin, C. Casaulta, C. Abbas, M. Bullo, O. Fuchs, E. Kieninger, I. Korten, L. Krüger, B. Seyfried, S. Yammine, and C. de Jong (University Children’s Hospital Bern, Bern, Switzerland); P. Iseli (Children’s Hospital Chur, Chur, Switzerland); K. Hoyler (private paediatric pulmonologist, Horgen, Switzerland); S. Blanchon, S. Guerin, and I. Rochat (University Children’s Hospital Lausanne, Lausanne, Switzerland); N. Regamey, M. Lurà, M. Hitzler, A. Clavuot, K. Hrup, and J. Stritt (Canton Hospital Lucerne, Lucerne, Switzerland); J. Barben (Children’s Hospital St Gallen, St Gallen, Switzerland); O. Sutter (private paediatric practice, Worb, Bern, Switzerland); A. Moeller, A. Hector, K. Heschl, A. Jung, T. Schürmann, L. Thanikkel, and J. Usemann (University Children’s Hospital Zurich, Zurich, Switzerland); and C.E. Kuehni, C. Ardura-Garcia, S. Glick, D. Berger, T. Krasnova, R. Makhoul, M. Ganbat, B. Guerra Buezo, F. Romero, M.C. Mallet, E.S.L. Pedersen, and M. Goutaki (University of Bern, Institute of Social and Preventive Medicine, Bern, Switzerland).

